# How do general practitioners consider health literacy in pain medication treatment of patients suffering from chronic musculoskeletal pain? *a mixed methods study*

**DOI:** 10.64898/2025.12.11.25341892

**Authors:** Rikke Bækgaard Nielsen, Kristian Damgaard Lyng, Alessandro Andreucci, Anne Estrup Olesen, Rasmus Østergaard Nielsen, Per Kallestrup, Michael Skovdal Rathleff

## Abstract

**Background:** Health literacy (HL) influences communication quality, treatment adherence, and equity in care. However, how general practitioners (GPs) recognize and respond to patients’ HL in everyday clinical reasoning remains insufficiently understood.

**Objective:** To investigate how Danish GPs incorporate patients’ health literacy into decisions about prescribing pain medication for chronic musculoskeletal pain, using insights from surveys, interviews, and a literature review.

**Methods:** A mixed-methods design combined survey data from 39 Danish GPs, seven qualitative interviews, and a synthesis of 14 studies on HL in general practice. The literature was used to contextualize and contrast the empirical findings. Quantitative data were analyzed descriptively, while qualitative data underwent thematic analysis. All three datasets were integrated through mixed-methods comparison to assess convergence, divergence, and complementarity.

**Results:** Across the integrated survey, interview, and literature findings, HL emerged as a largely implicit but consistent element of GP decision-making. In the Danish survey and interview data, some GPs explicitly reported considering HL in prescribing decisions, yet interviews showed that HL more often influenced clinical reasoning indirectly through intuition and conversation. GPs adapted communication, explanations, and treatment planning to their perceptions of patient understanding, but these adjustments were rarely guided by structured tools or frameworks. Conversation appeared as the main approach for assessing comprehension, echoing patterns observed in the literature. Many Danish GPs perceived most patients as competent and self-managing, a perception the literature cautions may mask hidden comprehension challenges. Finally, both local interviews and existing studies highlighted digital HL as an emerging theme, with GPs commonly managing patients’ online health information through conversational reframing rather than formal strategies.

**Conclusions:** HL is tacitly integrated into GP reasoning but remains under-recognized as a professional skill. Making HL an explicit component of communication training, reflective practice, and prescribing guidelines could improve patient understanding, shared decision-making, and treatment equity.

## Introduction

Chronic musculoskeletal (MSK) pain is one of the most common reasons for seeking medical care, with a significant proportion of primary care consultations related to acute or chronic pain conditions [1–3]. The influence of biological, psychological, and social factors undeniably makes pain management a challenging task that requires careful considerations of both clinical and patient-centered aspects [4–6]. Managing chronic pain in primary care has previously been recognized as one of the largest challenges faced by primary care physicians, [7]. The complexity of chronic pain in general also influences the patients, who must now navigate online platforms, websites, and apps for health information, heightening the need for skills to engage with the system effectively [8–10]. In response to these growing complexities, the concept of health literacy (HL) has gained increasing prominence in medicine and the health sciences. The World Health Organization (WHO) define health literacy as: “*representing the personal (health-related) knowledge and competencies that accumulate through daily activities, social interactions and across generations*”, and is linked to “*people’s knowledge, motivation, and competences to access, understand, appraise, and apply health information in order to make judgements and take decisions in everyday life concerning health care, disease prevention, and health promotion to maintain or improve quality of life during the life course*” [11].

People with chronic MSK pain are particularly vulnerable if they struggle with low HL, as this may impact their ability to self-manage their condition effectively [12–14]. Furthermore, research has shown that low levels of HL decrease self-confidence, adherence to treatment regimens and increases the risk of hospitalization and physical decline [15–18]. In Denmark, GPs serve as the first point of contact for these patients and act as gatekeepers to specialized healthcare services. GPs offer various treatment options, including analgesics, making their assessment and decision-making crucial to determining the patient’s treatment plan [19]. GPs’ treatment decisions not only affect individual patients but also have broader socioeconomic implications. Inappropriate referrals and ineffective treatment plans may prolong diagnostic processes and delay appropriate interventions, potentially increasing healthcare costs and reducing efficiency [20]. Low HL might hamper communication regarding symptoms and lived experiences, complicating GPs’ ability to deliver optimal care [21]. Because chronic MSK pain management largely relies on what patients do themselves, safe prescribing depends on patients’ HL, what GPs can expect them to understand and manage. However, it is unclear how GPs factor health literacy into their prescribing. Therefore, this study aims to investigate how Danish GPs consider patients’ health literacy when deciding on and communicating about pain medication for chronic MSK pain, drawing on evidence from surveys, interviews, and a literature review.

## Materials and Methods

### Study Design

The study adopted a pragmatic approach, which emphasizes the practical application of research to real-world problems and acknowledges that knowledge can be derived from both objective observations and subjective experiences. Pragmatism assumes that reality is complex and context-dependent, and it values the use of multiple methods to explore different dimensions of a research question. Rather than adhering strictly to a single philosophical tradition, the pragmatic approach focuses on using the most appropriate tools and strategies to answer the research question at hand. Hence, we employed a sequential explanatory mixed-methods design, integrating surveys, semi-structured interviews, and a literature review [22]. This approach was selected to obtain the benefits of several scientific paradigms and methods [23]. Firstly, we sought a broad overview of GPs’ clinical practices through a questionnaire followed by semi-structured interviews to explain the rationale behind their responses to the questionnaire. To add analytical depth in the analysis, we conducted a literature review focusing on HL research within pain research. Survey data were collected via an online survey hosted on REDCap™ (19). Qualitative data was collected via Microsoft Teams (Microsoft Corporation 2023. Microsoft Teams v.1.6), under COVID-19 restrictions. The study was conducted in accordance with the Helsinki Declaration and complied with European General Data Protection Regulation (GDPR) under approval from Center for General Practice at Aalborg University, Denmark (ID 131-2). In accordance with Danish ethical guidelines, the study did not require review by a national ethics committee, as no interventions were involved. However, informed digital consent was obtained from all participants prior to survey completion and interview participation. Reporting of the study followed the Good Reporting of A Mixed Methods Study (GRAMMS) [24].

### Participants and Recruitment

Danish GPs were recruited via advertisements distributed through professional networks (e.g., the Unit for General Practice in North Jutland Region, NordKAP) and social media platforms (e.g., a Facebook group for Danish doctors). An introduction letter outlining the study’s purpose and content accompanied all recruitment materials. GPs who consented to participate in the qualitative phase were invited for interviews via email, using the contact details provided in their response to the questionnaire. Data collection took place between March 14 and May 7, 2021. Participants received financial compensation according to standard reimbursement fees for general practitioners in Denmark.

### Development of the questionnaire

The survey included 11-item and three case vignettes to capture a range of data, including participant characteristics (e.g., age, gender, experience, information about their practice, special interests), type of decision for each case vignette (e.g., medication, exercise, referral), and the factors that influenced decisions to prescribe or withhold pain medication (e.g., patient needs, patient characteristics, presence of comorbidities) (the Danish survey and vignettes can be obtained by contacting the corresponding author). Responses were securely recorded through the REDCap™ platform.

### Development of interview guide

An interview guide (**Appendix 1**) was developed mainly based on the responses from the questionnaire and multi-professional expertise of the research group in the fields of pain management, general practice, and mixed methods. The interview guide underwent iterative revisions through research team discussions and was finalized following two pilot interviews with GPs. The interview guide explored GPs’ experiences treating chronic MSK pain patients in general practice, their preferred treatment strategies for chronic MSK pain, key factors influencing decisions to prescribe pain medication, and their perceptions of HL and its impact on decisions to prescribe medications. Questions were open-ended to facilitate an inductive approach, allowing the participants to share their perspectives freely. Written consent was obtained before initiation of the interview. All interviews were conducted in participants’ first language, with one exception. Each interview lasted 30-35 minutes and was video- and audio-recorded with participants’ consent for pseudonymized transcription. Data was securely stored at Aalborg University servers. Transcriptions were produced in a naturalized format, prioritizing meaning retention while minimizing nonverbal elements and speech irregularities to ensure an accurate reflection of participants’ statements.

### Literature review

To strengthen the analytical depth of survey and interview findings, a systematic literature search was conducted using a stepwise approach. This approach was used to accommodate the possibility of limited studies directly addressing the intersection of general practitioners, health literacy, and chronic MSK pain. This hierarchical strategy allowed us to begin with a narrow focus and gradually broaden the scope to ensure comprehensive coverage of the literature. All searches were performed in PubMed, Embase, and PsycINFO, covering studies published up to July 2025. The literature search was conducted in three consecutive steps, gradually expanding the scope to ensure comprehensive coverage. Step 1 focused on studies specifically examining health literacy in the context of chronic MSK pain and pain medication within general practice. Step 2 broadened the search to include studies addressing HL and chronic MSK pain in general practice, regardless of treatment type. Step 3 further extended the scope to encompass all studies investigating HL in general practice, independent of condition or treatment focus. Across all three steps, studies were included if they investigated adult populations and that examined health literacy or closely related constructs, e.g. patient understanding, communication, or condition-specific knowledge, within the context of general practice. We excluded studies involving participants under 18 years of age, studies that focused solely on general literacy rather than health literacy, and studies concerned primarily with validating measurement instruments. In addition, we excluded studies conducted in healthcare systems not comparable to the Danish context, as well as studies whose primary focus was on mental health conditions. Search strategies were adapted for each database (See full search strings in **Appendix 2**).

### Data analysis Quantitative analysis

Descriptive statistics were calculated to summarize GPs characteristics, including demographics, experience, practice size and type, special interest in chronic MSK pain, and typical treatment choices. Differences between GPs who considered patients’ HL in treatment decisions and those who did not were analyzed using independent t-tests for continuous variables and chi-square or Fisher’s exact tests for categorical variables. All statistical analyses were conducted using SPSS version 28.0, with a significance level set at *P* < 0.05.

### Qualitative analysis

The data were analyzed using NVIVO v. 13, following a reflexive thematic analysis as outlined by Braun and Clarke [25]. The six phases included: (1) familiarization with the data, (2) generating initial codes, (3) constructing initial themes, (4) reviewing themes, (5) defining and naming themes, and (6) reporting. An inductive approach was used, allowing themes to be generated from the data rather than from pre-existing frameworks. Consistent with the reflexive approach, the researcher (RBN) actively engaged with the data and her own subjectivity throughout the analytic process. To ensure consistency and depth of interpretation, all interviews were conducted, transcribed, translated, and analyzed by the same researcher.

### Literature review

The results of the literature search were screened in two stages: title/abstract screening followed by full-text review, based on the predefined eligibility criteria. Studies meeting the inclusion criteria were imported into Covidence, a web-based collaboration software that assists the production of systematic and other literature reviews (Covidence systematic review software, Veritas Health Innovation, Melbourne, Australia. Available at www.covidence.org) and duplicates were removed. For each included study, we extracted the following information: study design, country and healthcare setting, participant characteristics, conceptualization and measurement of HL, and key outcomes related to answering the aim of our study. Extracted data were synthesized narratively to identify themes and knowledge gaps relevant to the role of GPs in addressing HL among patients with chronic MSK pain. All steps were conducted by two authors (RBN and KDL).

### Mixed Methods Data Integration

Integration was conducted at two stages of the study. First, data from the survey were used to inform the development of the interview guide (connection). Second, following the completion of the survey, interviews, and literature review, findings were integrated through a mixed-methods analysis. This analytical integration involved a juxtaposed comparison of the three datasets to assess areas of confirmation (overlapping findings), expansion (complementary or elaborating insights), and discordance (divergent or contradictory results) [23,26]. This comparative process enabled the development of meta-inferences (interpretations derived from synthesizing evidence across quantitative, qualitative, and literature-derived data) to generate a deeper and more contextualized understanding of how health literacy influenced GPs’ decision-making in relation to pain medication treatment. Integration at the reporting level was achieved through weaving, where survey, interview, and literature findings were presented together under shared thematic headings. Finally, a joint display was used to visually organize and communicate the integrated results, facilitating transparency in how inferences were drawn across the three data strands [27,28].

## Results Demographics

### Survey and interview participation

A total of 39 GPs completed the questionnaire. Among the 39 general practitioners, 12 initially indicated willingness to participate in the subsequent interviews, but due to scheduling conflicts, only five interviews were conducted. An additional two GPs were recruited through the research network, bringing the final qualitative sample to seven GPs. **Table 1** summarizes the demographic and professional characteristics of the survey and interview participants.

**Table 1.** Participant Demographics.

|  | Survey | Interview |
| --- | --- | --- |
|  | N = 39 (%) | N = 7 (%) |
| **Sex** |  |  |
| Male: | 17 (43,59) | 5 |
| Female: | 22 (56,41) | 2 |
| **Age (years)** |  |  |
| Mean: | 44,12 | - |
| SD: | ± 6,31 | - |
| Min: | 34 | 34 |
| Max: | 64 | 64 |
| **Years of experience** |  |  |
| 1-5 years | 16 (41,03) | 2 |
| 6-10 years | 14 (35,90) | 1 |
| 11-15 years | 6 (15,38) | 1 |
| 16-20 years | 1 (2,56) |  |
| 21-25 years | 1 (2,56) | 2 |
| 26-30 years | 1 (2,56) | 1 |
| **Practice size** |  |  |
| <2000 patients | 7 (17,95) |  |
| 2000-4000 patients | 8 (20,51) | 1 |
| 4000-6000 patients | 12 (30,77) | 4 |
| >6000 patients | 12 (30,77) | 2 |
| **Practice location** |  |  |
| >10000 inhabitants | 27 (69,23) | 6 |
| 5000-10000 inhabitants | 5 (12,82) | 1 |
| <5000 inhabitants | 7 (17,95) |  |
| **Type of practice** |  |  |
| Solo practice (not part of a collaborative) | 3 (7,69) | 1 |
| Solo practice (part of a collaborative) | 4 (10,26) | 2 |
| Company practice | 32 (82,05) | 4 |
| **Special interest** |  |  |
| Yes | 9 (23,08) | 1 |
| No | 27 (69,23) | 6 |
| Don't know | 3 (7,69) |  |

### Quantitative results

Of the 39 GPs surveyed, only 3 (8%) reported primarily prescribing painkillers for MSK pain, while 27 (69%) used a mixed strategy combining medication with other treatments. Twenty-three GPs (59%) emphasized tailoring treatment for the individual patient. The most commonly reported factors influencing prescribing decisions were pain characteristics and comorbidities (both 76.9%), followed by knowledge of the patient (69.2%), medical history and medication effects/side effects (both 64.1%), and concurrent medication use (61.5%). Broader patient characteristics (56.4%), daily needs (43.6%), and lack of alternative strategies (33.3%) were less frequently cited. Only 5 GPs (12.8%) mentioned the availability of clinical guidelines. Of the 39 GPs, 38.5% considered a patient’s HL level relevant when prescribing pain medication, and the same proportion emphasized the patient’s ability to self-care. Fewer (28.2%) regarded the ability to understand health information as important (See **Figure 2**).

**Figure 1:**
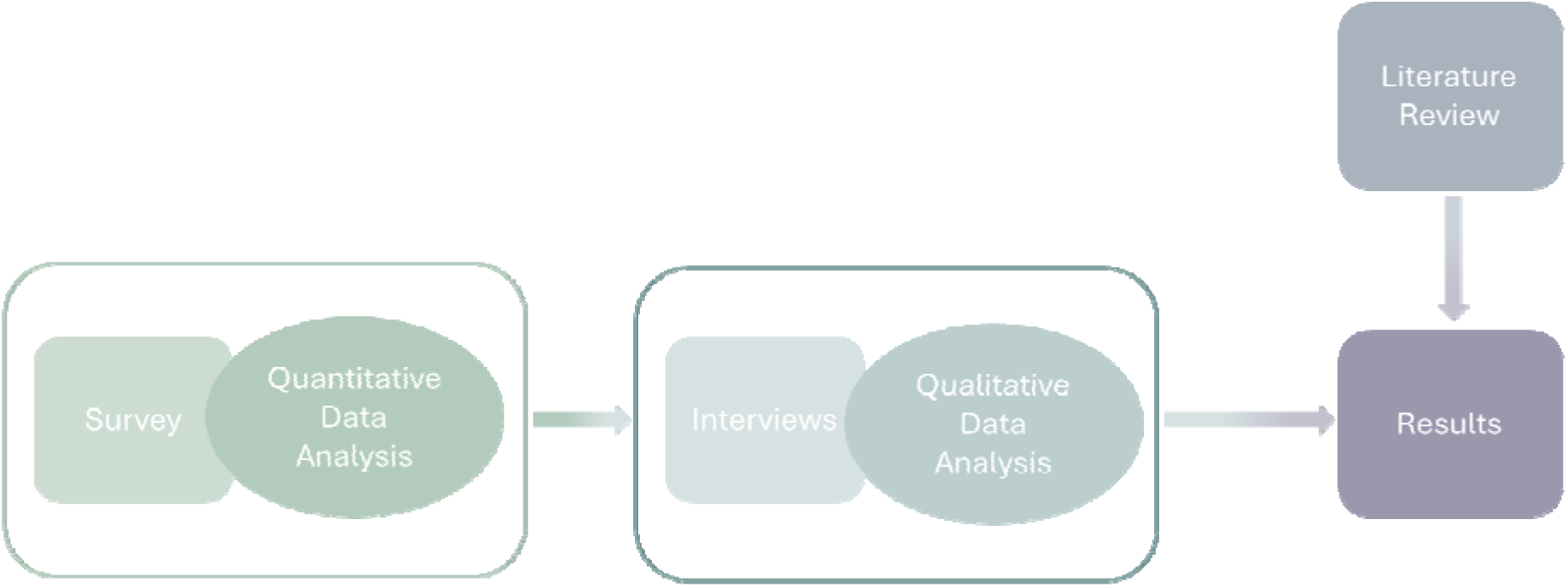
Visualization of the study process.

**Figure 2.**
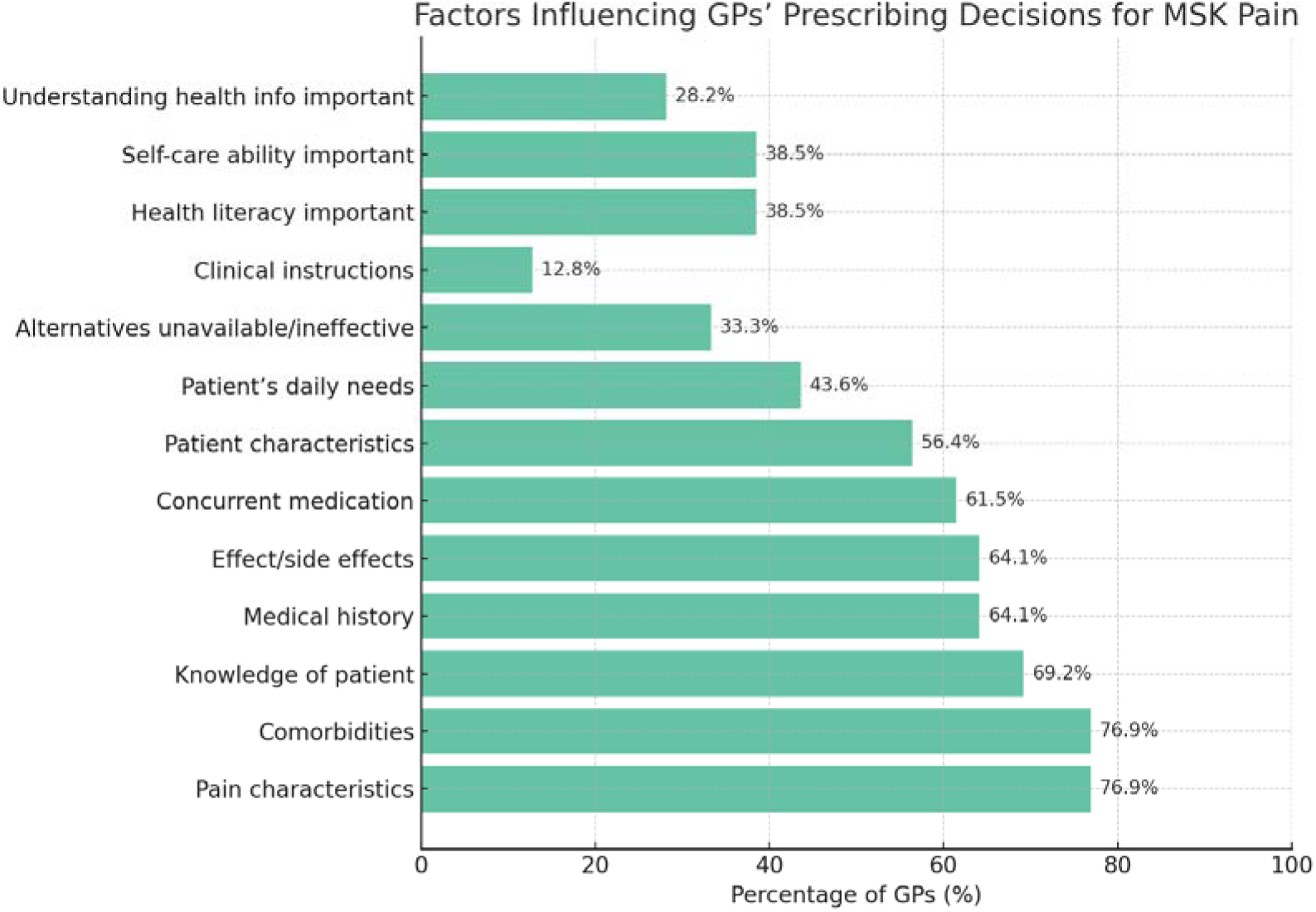
Factors influencing GPs prescribing decisions for chronic MSK pain.

Independent t-tests showed that GPs who considered HL did not differ significantly from those who did not in terms of age (p = 0.91). Fisher’s exact tests revealed no significant associations with gender or special interest in MSK pain (both p > 0.70). Due to the small sample size and sparse data across several categories, other variables such as years of experience and practice size were not included in formal statistical comparisons. Attempts to identify independent predictors of HL consideration using logistic regression were also unsuccessful, as the models failed to converge despite variable simplification.

### Qualitative Results

The qualitative part of the project identified three main themes, with one subtheme. It expanded the understanding of the survey results, in particular the finding of less than a third of the GPs deeming health information important. The seven interviews revealed that while HL was rarely named explicitly by GPs, it played a subtle yet consistent role in their clinical reasoning and patient communication. HL appeared to be an intuitive consideration, embedded in how GPs assessed understanding, tailored information, and supported patient decision-making – often without labeling these actions as related to HL.

#### Theme 1: Automated inclusion of health literacy

While only one GP explicitly identified HL as a decisive factor in their clinical reasoning, the majority of participants demonstrated that HL was nevertheless embedded in their everyday practice. Rather than being treated as a standalone concept, HL seemed to function as an invisible competency – something assessed and acted upon intuitively rather than consciously. GPs described adapting their explanations, gauging patient comprehension, and tailoring treatment strategies based on how well patients seemed to grasp and apply health-related information.

As one GP succinctly expressed:

> “I’ll say that is something you do kind of automatically.”

> — General Practitioner 3

Even in responses to questions unrelated to HL, participants revealed how communication and patient understanding influenced their clinical decisions. For instance, one GP linked professional experience not only to improved treatment planning but also to the development of patient education skills:

> “The more experienced you are, the better you become at making a treatment plan […] and at teaching the patients how to take the medicine and what to pay attention to.”

> — General Practitioner 1

This suggests that HL is not always consciously named or prioritized, but rather operates as an embedded aspect of competent, relational clinical care – particularly for GPs with years of experiential learning.

#### Theme 2: Conversation as a tool

Conversation was consistently described as the central mechanism through which GPs assessed, supported, and responded to patients’ HL. Rather than relying on formal assessment tools or standardized questions, GPs drew on conversational cues to determine whether patients understood their condition, the proposed treatment, and their role in managing it. GPs described repeating key points, asking open-ended questions to confirm comprehension, and summarizing instructions both verbally and in writing. These communicative strategies were not specific to patients with MSK pain but were viewed as part of good general practice:

> “I always ask the patient if they understand what I have said, and I often write a little note, which I hand out to the patient – a kind of plan. I do this generally, not only with pain patients.”

> — General Practitioner 3

Conversation also allowed for flexibility and personalization – two features that may be especially important when addressing patients’ varying levels of HL. The GPs’ responses reflected a pragmatic and patient-centered approach, where the consultation served not just as a diagnostic tool, but also as an educational space.

### Subtheme: “Dr. Google”

The GPs acknowledged that many patients now engage with health information online before seeking professional care. This phenomenon – sometimes referenced as “Dr. Google” – was viewed with a mixture of tolerance and caution. While GPs recognized that most patients seek online information in good faith but explained that problems may arise when this information is misunderstood or applied inappropriately. This was described as particularly challenging in the diagnostic phase, when patients may arrive with fixed ideas about their condition based on self-diagnosis.

> “People come in and are completely certain that they suffer from something, but in reality, it’s probably just because they didn’t entirely understand what they read.”

> — General Practitioner 7

Despite these challenges, GPs generally approached such situations constructively, using conversation, diagrams, or models to reframe misinformation and re-establish shared understanding. This subtheme suggests the presence of a dynamic tension between patient agency and the risk of misinformation and underscores the importance of communication skills in navigating HL in the digital age.

#### Theme 3: Skillful patients

A recurring narrative among GPs was that most patients are already competent in managing their symptoms and making basic treatment decisions before consulting their doctor. Participants frequently described patients as informed, proactive, and resourceful, qualities closely aligned with functional HL, even if not labeled as such. This was expressed for example in these two statements:

> “By far most people are actually relatively good at taking care of themselves and react, so I don’t have real problems with [health literacy].”

> — General Practitioner 1

> “Most of them have at least tried taking paracetamol. Some [patients] started NSAIDs before coming.”

> — General Practitioner 6

This theme suggests that GPs may underestimate the need to explicitly consider HL because they perceive their patient population as already possessing adequate knowledge and skills. It also points to an important contextual nuance: HL is not merely a matter of patient deficits but also of patient capabilities, which GPs often recognize and rely upon in clinical encounters.

#### Literature review

The literature search identified 13.077 individual hits of which 11.828 were screened through title and abstract. Then 364 studies were screened for eligibility based on full-text and finally, 14 papers were included for further analysis (See **figure 3**).

**Figure 3.**
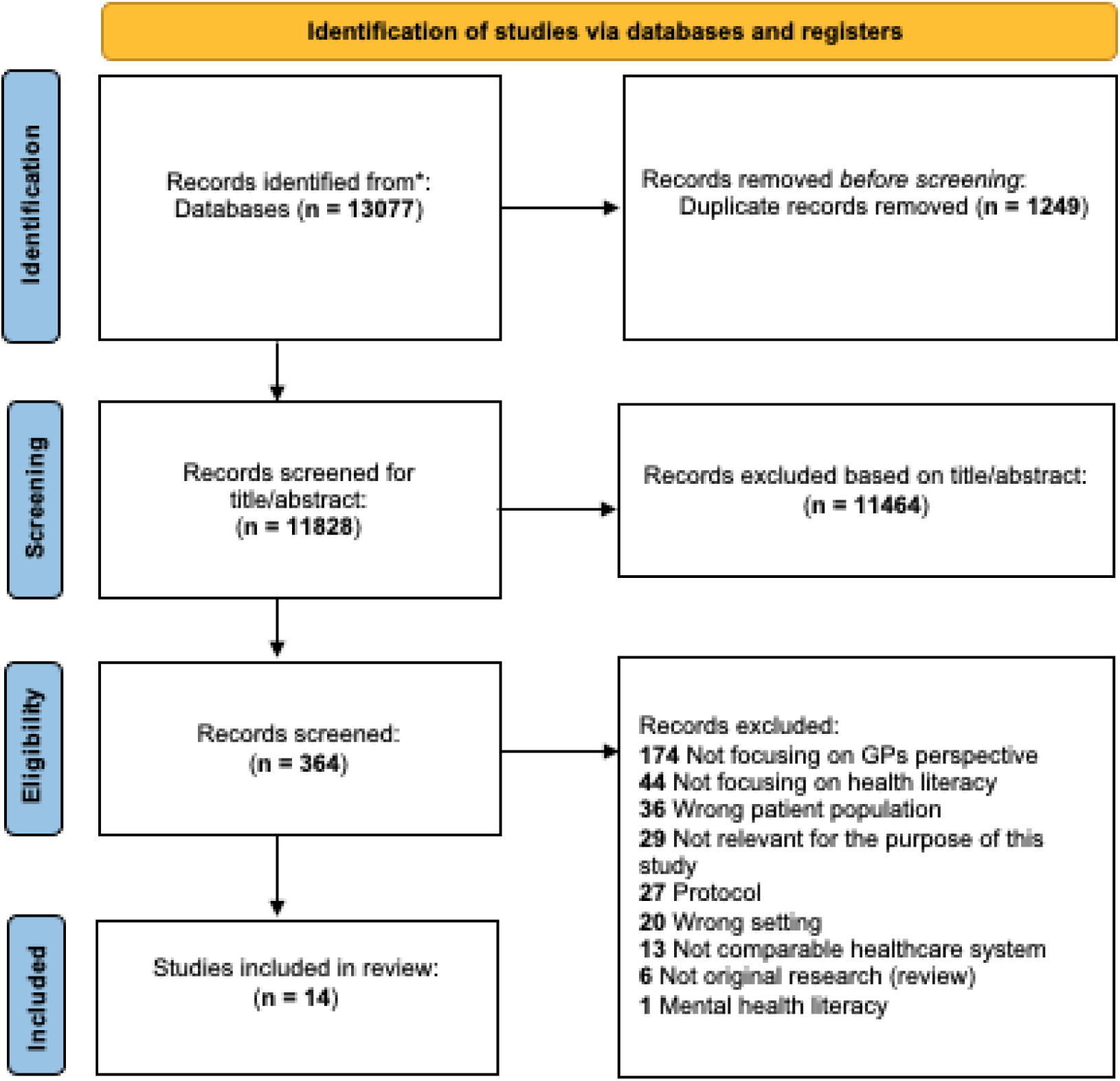
Flowchart of the literature search.

### Characteristics of the included studies

The 14 studies involved 656 GPs across Australia (3 studies, 21%; 284 GPs, 43%), the US (5, 36%; 153 GPs, 23%), and Europe (6, 43%; 219 GPs, 33%) – plus 2,824 patients and 49 other health professionals. Most focused on chronic disease management, particularly diabetes (4 studies, 29%), followed by cancer screening (2, 14%), multimorbidity (2, 14%), and polypharmacy or cardiovascular prevention (2, 14%). Four studies (29%) examined system or digital literacy. Study designs included qualitative (6, 43%), surveys (4, 29%), mixed methods (2, 14%), and RCTs (2, 14%). Ten studies used validated HL tools, mainly Test of Functional Health Literacy in Adults (TOFHLA), Rapid Estimate of Adult Literacy in Medicine (REALM), or The Newest Vital Sign (NVS) in US settings and HLS-EU-Q16 in Europe. Most assessed functional literacy (10, 71%), with fewer addressing interactive/critical (4, 29%) or digital literacy (3, 21%). Several qualitative studies viewed HL as a relational practice within communication and decision-making.

### Narrative synthesis of literature review

Across the included studies, HL consistently emerged as a key determinant of patient engagement, communication quality, and treatment management in primary care and chronic disease contexts. Several studies demonstrated that GPs often overestimate patients’ HL, leading to mismatched communication and limited shared decision-making. Kelly and Haidet (2007) found that physicians overestimated patient literacy in more than half of encounters, particularly among minority patients [29], a finding corroborated by Storms et al. (2019), who reported only slight agreement (κ = 0.033) between Belgian GPs’ HL estimates and patients’ self-reported HL [30]. Similarly, McKinn et al. (2024) showed that Australian GPs rely on subjective impressions rather than structured assessments, adjusting communication style based on perceived HL but without formal tools, potentially reinforcing inequities in preventive care [31]. Studies focusing on specific conditions further highlighted the clinical impact of limited HL. Schillinger et al. (2003), Fransen et al. (2014), and Seligman et al. (2005) demonstrated that in diabetes care, patients with low HL experience poorer understanding and glycemic control, while physician communication strategies such as “closing the loop” and explicit comprehension checks were strongly associated with better outcomes [32–34]. However, even when physicians were informed of their patients’ low HL, changes in clinical practice were modest without additional training or systemic support [33]. Research on patient experiences echoed these communication challenges. Gunn et al. (2020) found that women with limited HL desired shared decision-making around mammography but lacked technical and process knowledge, while primary care providers often avoided detailed discussions for fear of confusion [35]. Hamrosi et al. (2013) showed similar tensions in medication communication, where Australian GPs and pharmacists hesitated to provide written information due to perceived patient misunderstanding, despite strong patient demand for such materials [36]. Van Eikenhorst et al. (2019) and Herzig et al. (2019) both emphasized that patients’ self-management capacity depends not only on HL but also on trust, relational continuity, and psychosocial context, factors often underappreciated by clinicians, who tend to focus on biomedical complexity rather than treatment burden or comprehension barriers [37,38]. At the system level, Hailu et al. (2024) illustrated how digital and health literacy jointly shape the adoption of remote patient monitoring in primary care [39]. Low HL and digital skills among older adults constrained engagement, underscoring the intersection of literacy domains in modern healthcare delivery. Davidge et al. (2023) similarly showed that patients’ varying digital literacy levels influence the success of electronic health record access initiatives in primary care, with staff highlighting both empowerment opportunities and communication challenges [40]. Carzorla et al. (2025) and Gillespie et al. (2023) similarly pointed to HL as central to implementation success, particularly in deprived areas or among older adults managing polypharmacy [41,42]. Collectively, these studies reveal a persistent gap between the ideal of patient-centered communication and everyday primary care practice. HL remains insufficiently assessed and inconsistently addressed, with both patients and clinicians expressing a need for practical tools, pre-visit education, and system-level strategies to support equitable understanding, self-management, and shared decision-making.

### Results of mixed methods data integration

Taken together, the survey, interviews, and literature highlight that health literacy is present but largely unacknowledged in GP decision-making, often inferred intuitively rather than assessed or acted upon systematically. Quantitative results showed that biomedical factors such as pain characteristics and comorbidities dominated decision-making, with HL-related considerations mentioned far less frequently. In contrast, the qualitative findings revealed HL as an embedded but rarely explicit element of practice, expressed through intuitive judgments, conversational cues, and perceptions of patient competence. The literature echoed this pattern, showing that GPs often overestimate patient HL, rely on proxies rather than structured tools, and underutilize proven strategies such as teach-back. The following joint displays illustrate how the quantitative, qualitative, and literature findings align, diverge, or extend one another, offering a more comprehensive understanding of how GPs consider HL in prescribing decisions for chronic MSK pain [26,28]. Table 3 (below) presents a joint display integrating findings from the quantitative survey (n = 39 GPs), qualitative interviews (n = 7 GPs), and the literature review (14 studies).

**TABLE 2:**
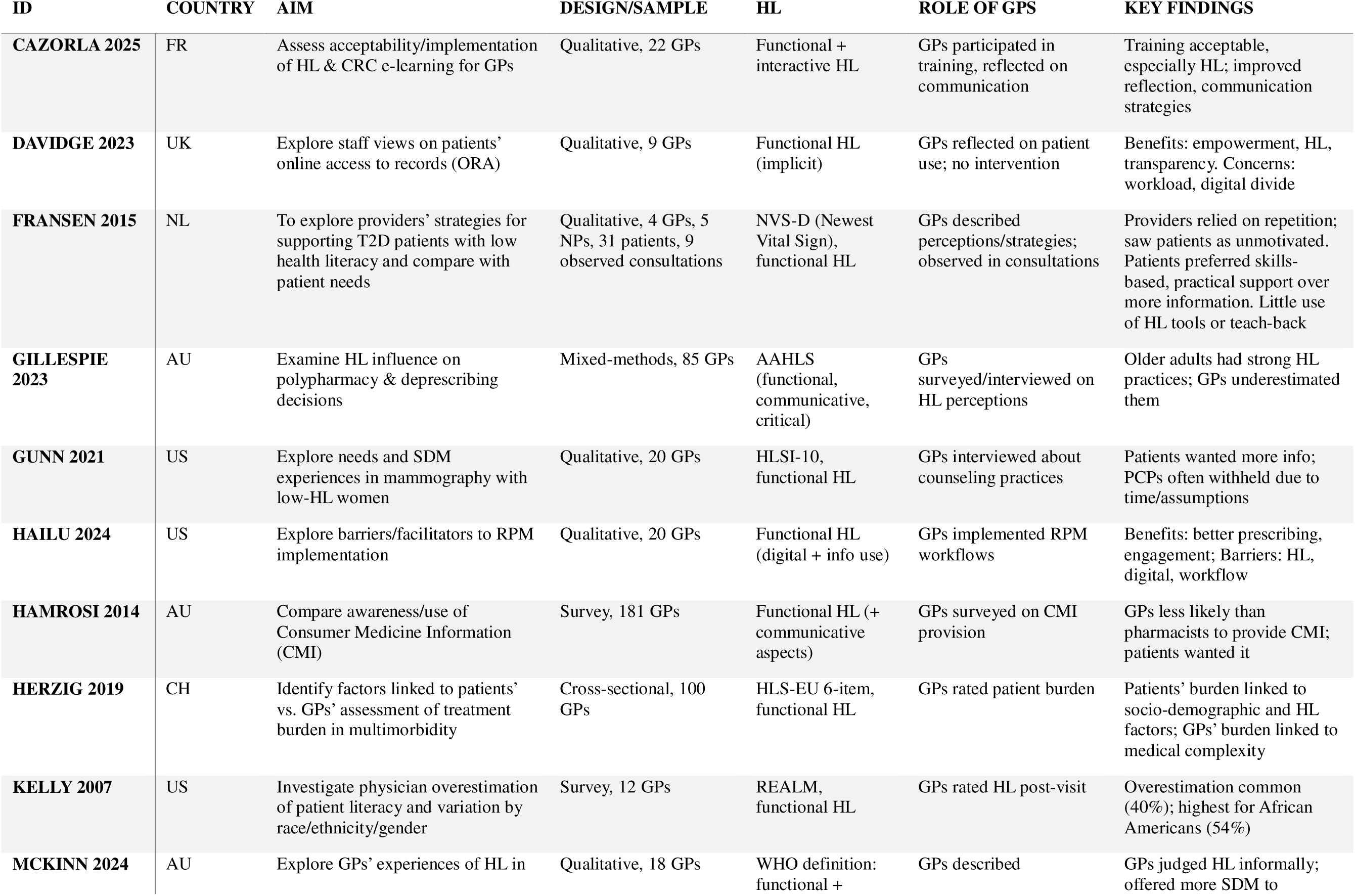
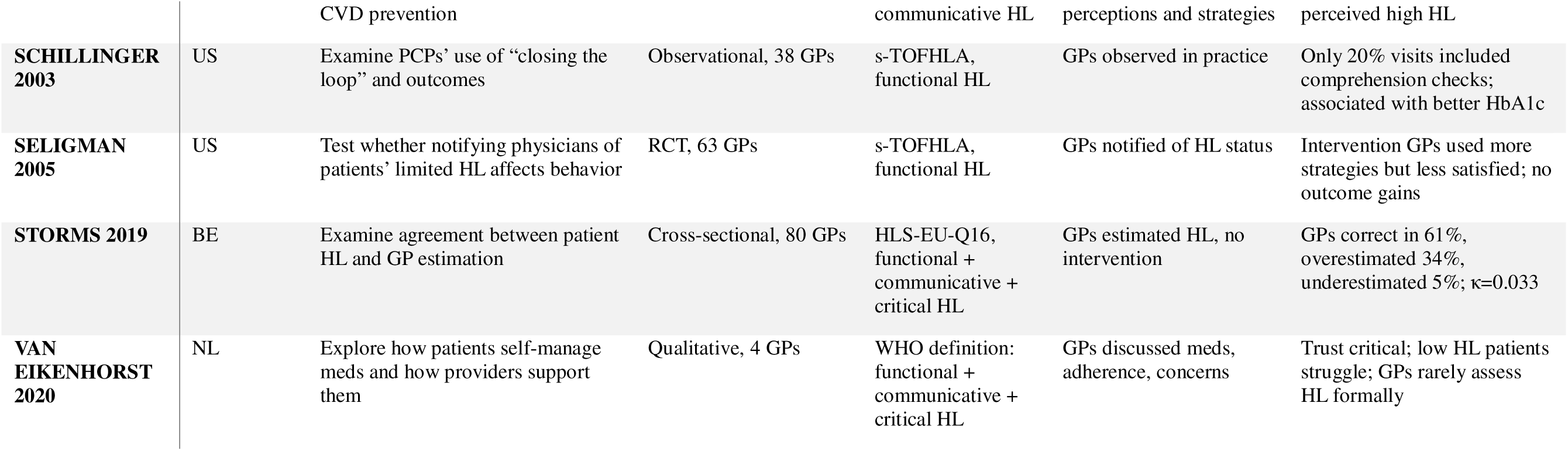
Study Characteristics.

**Table 3:** Joint display of quantitative and qualitative findings, as well as findings from literature.

|  | DATA TYPE |  |  | INTEGRATION LEVEL |
| --- | --- | --- | --- | --- |
| Focus area | Quantitative survey<br>(n=39 GPs) | Qualitative interviews<br>(n=7 GPs) | Literature<br>(14 studies) |  |
| **Explicit use of HL** | 38.5% considered HL important; 38.5% emphasized self-care; 28.2% emphasized understanding health information | HL rarely named explicitly, but embedded in reasoning as an “invisible competency” (Theme 1) | GPs often overestimate HL, rely on intuition rather than validated tools<br>(Kelly 2007; Storms 2019; McKinn 2024) | **Divergence:** Quantitative suggests low explicit use, while qualitative and literature show HL as implicit but influential |
| **Assessment of patient understanding** | Not systematically reported; main focus on pain characteristics (76.9%), comorbidities (76.9%), and medical history (64.1%) | Conversation described as the main tool: repetition, open-ended questions, written notes (Theme 2) | Formal HL assessment rare; teach-back seldom used but improves outcomes<br>(Schillinger 2003) | **Complementarity:** Survey highlights biomedical focus; qualitative and literature add mechanisms of communication |
| **Influence of patient context** | Knowledge of patient (69.2%), patient’s daily needs (43.6%) considered relevant | Patients viewed as “skillful” and proactive in self-care (Theme 3) | Patients show HL in everyday management, but GPs underestimate or overlook this<br>(Gillespie 2023; Fransen 2015) | **Convergence:** All strands emphasize patient context, though literature shows more underestimation than our interviews |
| **Digital health literacy** | Not captured in survey variables | Patients often bring online info (“Dr. Google”), creating both opportunities and challenges (Subtheme) | Patients increasingly use online information; concerns about misinterpretation and workload for GPs<br>(Davidge 2023; Hailu 2024) | **Expansion:** Emerged only qualitatively and in literature, not surveyed quantitatively |
| Communication strategies | Side effects and expected efficacy considered by 64.1%; limited mention of HL-specific strategies | GPs adapt communication pragmatically, without labeling it as HL-sensitive | Training interventions acceptable but rare; awareness alone insufficient<br>(Seligman 2005; Cazorla 2025) | **Convergence/Complementarity:**<br>All strands show limited explicit HL strategies; literature adds evidence on intervention outcomes |

## Discussion

### Summary of findings

This study investigated how Danish GPs consider HL when prescribing pain medication for patients with chronic MSK pain. While some surveyed GPs explicitly reported considering HL in prescribing decisions, interviews revealed that HL plays an implicit and consistent role in clinical reasoning. GPs frequently adapted communication, explanations, and treatment planning to how they perceived patients’ understanding, but these adjustments were largely intuitive rather than informed by structured tools or frameworks. Conversation emerged as the main means of assessing and supporting patient understanding. Many GPs perceived most patients as competent and self-managing, although this perception risked overlooking those with hidden comprehension challenges. Finally, digital HL surfaced as an increasingly important theme, with GPs describing encounters shaped by online health information and typically managing this through conversational reframing rather than formal strategies.

### Interpretation of results and comparison with existing literature

Our findings show that HL was rarely treated as an explicit clinical consideration but surfaced implicitly through relational and communicative practices. HL functioned less as a measurable skill and more as a tacit element of professional intuition, determined by cues sensed in conversation rather than assessed systematically. This mirrors international evidence that clinicians often rely on subjective judgments of patients’ understanding, yet our data add nuance by illustrating how such intuition is enacted in practice [29–31]. Danish GPs described “reading” patients through tone, confidence, and engagement. In this way, HL became intertwined with broader moral and relational assessments rather than a specific communicative domain, consistent with research showing that HL awareness is embedded within professional empathy rather than structured as a formal competence [37,38]. However, the tendency to interpret apparent confidence as sufficient understanding echoes concerns raised in previous literature, cautioning that engagement can mask significant comprehension gaps [29,33]. This suggests that intuitive communication, though relationally sensitive, risks reinforcing hidden inequities if not accompanied by tools or training that make HL more visible and actionable [41]. In the Danish context, where a substantial proportion of adults report difficulties in understanding or applying health information, relying solely on intuition becomes particularly complex [10,43,44]. While intuitive judgement enables GPs to adapt dynamically to patients’ needs, it also introduces uncertainty and the risk of misinterpreting patients’ actual comprehension. In our study, GPs frequently used dialogue as a corrective tool, e.g. listening for subtle signs of misunderstanding, reframing information, and re-establishing shared meaning. This adaptive approach echoes previous research demonstrating that strategies such as “closing the loop” and explicitly checking comprehension improve patient outcomes [32,35,36]. These findings underscore that HL is co-constructed in real time through communication rather than simply possessed or lacking, highlighting why implicit approaches alone may be insufficient for equitable prescribing in chronic MSK pain. Importantly, our findings extend beyond communication alone to show how HL shaped concrete prescribing decisions. GPs reported that their perceptions of a patient’s understanding and “resourcefulness” influenced how much medication complexity they felt comfortable prescribing, whether to escalate, de-escalate, or modify treatment, and how extensively they explained risks such as dependence, side effects, and interactions. When patients appeared verbally fluent or confident, GPs often assumed they could manage complex regimens or monitor for safety concerns, yet this confidence did not always reflect actual comprehension. These dynamics align with studies showing that unrecognized HL challenges can affect medication adherence, safety monitoring, and understanding of dosing instructions [30,36]. Thus, HL may shape not only the conversation about treatment but also the treatment decisions themselves. Our findings also reflect the shifting digital landscape that has transformed how patients engage with health information since data collection in 2021. At that time, patients drew primarily on “Dr. Google,” similar to patterns reported by van Riel et al. 2017 [45] and Luo et al. 2022, where online information shaped but sometimes strained doctor–patient dialogue [46]. GPs sought to re-anchor these conversations in shared understanding rather than dismiss online input, consistent with newer research on digital communication and patient empowerment [40,47]. Since 2021, however, the information environment has evolved rapidly. Generative AI now produces fluent, personalized, and persuasive health content, shifting the challenge from information overload to epistemic competition between clinician advice and algorithmic explanations [48,49]. As a recent study from Comeau et al. 2025 argues, interpretation of health information is now performed not only by patients but also by algorithms, further complicating the communicative and relational foundation on which safe prescribing rests [50].

### Strengths and Limitations

A main strength of this study is the mixed-methods design, which allowed for depth in exploring how GPs consider HL when managing patients with chronic MSK pain. Combining survey, interview, and literature data enabled triangulation and strengthened analytical interpretation. Conducting all phases within a pragmatic framework ensured that the findings remained closely connected to clinical practice. A number of limitations should be noted. The sample was small and potentially non-representative, as recruitment through professional networks and social media may have led to self-selection of GPs with a particular interest in pain management or HL. The limited number of respondents reduced statistical power, preventing meaningful regression or subgroup analyses. Recruiting GPs for survey and interview studies in Denmark is consistently challenging due to heavy time pressures and limited availability for research activities; thus, large sample sizes are rarely feasible, and the achieved response rate is high for this population. Data collection took place during the COVID-19 pandemic, though this context is unlikely to have influenced the findings. Conducting, transcribing, and analyzing all interviews by one researcher ensured internal consistency but introduces potential interpretive bias despite ongoing reflexivity. Translation from Danish to English may have led to minor loss of nuance in expressions related to communication or HL. Integrating three datasets was methodologically demanding; while genuine synthesis was sought, some degree of parallel reporting may remain.

### Implications

At the clinical level, our findings suggest that HL is already woven into GPs’ communication and prescribing practices, though it remains largely implicit. Making HL an explicit skill, e.g. through awareness, structured communication training, and use of techniques such as teach-back, could reduce misunderstandings and promote equity, particularly in prescribing decisions for chronic MSK pain. Explicit HL practices could support safer dosing, clearer monitoring of risks, and better alignment of expectations in long-term treatment. At the organizational level, our results support HL as a shared system responsibility rather than an individual trait, aligning with WHO policies [51]. Integrating HL principles into education, guidelines, and everyday workflows could reduce reliance on intuition and make prescribing conversations more consistent and equitable. As patients increasingly use digital and AI-generated content, health tools and communication strategies should be designed to be transparent, comprehensible, and accessible for diverse patient groups. Continued attention to both relational and digital HL will be essential for maintaining shared understanding, supporting safe prescribing, and sustaining trust in primary care.

### Conclusion

This study shows that HL is present but mostly implicit in Danish GPs’ prescribing for chronic MSK pain. While surveys highlight biomedical drivers, interviews reveal HL is handled intuitively through conversation, informal checks, and reframing patients’ online beliefs, sometimes overestimating patient competence and masking gaps. Making HL explicit could improve equity and safety: use plain language, brief teach-back, and written medication plans as routine “universal precautions,” and embed HL in education and workflows so it becomes a shared system responsibility. As digital and AI-generated information increasingly shapes patient views, tools should be clear, transparent, and accessible. Future research should therefore examine how HL can be systematically assessed and operationalized in real-world consultations, and how structured HL-supportive practices can be sustainably integrated into GP workflows.

## Data Availability

All data produced in the present study are available upon reasonable request to the authors

## Declarations

### Consent for publication

All authors have read and approved the final manuscript and consent to its publication.

### Availability of data and materials

Data are available upon request to authors and in supplementary materials.

### Authors’ contributions

*Conceptualization:* Rikke Bækgaard Nielsen, Kristian Damgaard Lyng, Alessandro Andreucci, Anne Estrup Olesen, Rasmus Østergaard Nielsen, Per Kallestrup, Michael Skovdal Rathleff

*Methodology:* Rikke Bækgaard Nielsen, Alessandro Andreucci, Kristian Damgaard Lyng, Michael Skovdal Rathleff.

*Software*: Rikke Bækgaard Nielsen, Alessandro Andreucci, Kristian Damgaard Lyng.

*Validation*: Rikke Bækgaard Nielsen, Kristian Damgaard Lyng, Alessandro Andreucci, Anne Estrup Olesen, Rasmus Østergaard Nielsen, Per Kallestrup, Michael Skovdal Rathleff.

*Formal analysis:* Rikke Bækgaard Nielsen, Kristian Damgaard Lyng, Alessandro Andreucci,

*Investigation*: Rikke Bækgaard Nielsen, Kristian Damgaard Lyng, Alessandro Andreucci.

*Resources:* Rikke Bækgaard Nielsen, Kristian Damgaard Lyng, Alessandro Andreucci, Michael Skovdal Rathleff.

*Data curation:* Rikke Bækgaard Nielsen, Kristian Damgaard Lyng, Alessandro Andreucci.

*Writing – Original Draft:* Rikke Bækgaard Nielsen, Kristian Damgaard Lyng.

*Writing – Review & Editing:* Alessandro Andreucci, Anne Estrup Olesen, Rasmus Østergaard Nielsen, Per Kallestrup, Michael Skovdal Rathleff.

*Visualization*: Rikke Bækgaard Nielsen, Kristian Damgaard Lyng.

*Supervision:* Anne Estrup Olesen, Rasmus Østergaard Nielsen, Per Kallestrup, Michael Skovdal Rathleff.

*Project administration*: Rikke Bækgaard Nielsen, Kristian Damgaard Lyng, Michael Skovdal Rathleff.

*Funding acquisition*: N/A.

