## Appendix 1 for "How do general practitioners consider health literacy in pain medication treatment of patients suffering from chronic musculoskeletal pain? *a mixed methods study*"

### Appendix 1. Interview guide

| Purpose | Theme | Questions | Follow-up | Comments |
| --- | --- | --- | --- | --- |
| Opening |  | Hi. My name is Rikke, im from AAU. You have been emailing with my colleague, about our study about the use of pain meds.  Consent:  Is it okay that i record the interview? (You will be anonymous in the study and data will be stores cf. GDPR). | They can ask if they have questions. |  |
| “Icebreaker” + Leading their thoughts to pain treatment at their job | MSK patients in general | What is the typical patient with MSK complaint you see in your clinic? | - Do you see more of those than other diagnoses?  what are the typical types of treatments you might use with patients? |  |
| Icebreaker bridge | Pain medication in general | How do you see the role of pain medication for MSK pain? | Which types of pain medication?  When?  Is it often something you use early on? |  |
| Follow up page 9 of questionnaire (Generally) | Important factors | From your experience, what are the most important factors to consider when prescribing pain medication? | -Why do you consider these factors as the most important?  - Are there any other important factors that should be investigated and were not covered in the survey?  - From your experience, what is the greatest issue when deciding ***to*** prescribe or ***not*** prescribe pain medication? |  |
| Bridge from open questions to specific questions | Pain medication in general | Did your attitude regarding the prescription of pain medication change throughout years? | If yes, why? And how do you generally feel about pain meds and prescribing them?  If no, how do you generally feel about pain meds and prescribing them? |  |
| Follow up Patient daily needs and Patient characteristics (from page 9) | Patient characteristics | Can you describe a situation where factors like **age, gender, job** or alike, led you ***to*** prescribing pain med. and a situation where these factors led you to ***not*** prescribe pain meds? | What about that situation/factor led you to make one decision or the other? |  |
| Follow up Pain characteristics (from page 9) | Pain characteristics | Another factor that might play a part is the **pain characteristics** (e.g. pain severity, frequency, duration location) how does these factors affect your choice to prescribe pain medication? | Can you describe a situation where you decided ***to*** prescribe pain medication and a situation where you decided ***not*** to prescribe pain medication due to the pain characteristics?  Are there pain characteristics that are more likely to influence your decision than others? |  |
| Follow up Effects and side effects (from page 9) | Effects and side effects | The effects of pain medication and the side effects might influence the decision to prescribe pain medication. **How does the expected effect of pain medication and potential side-effects, affect your choice to prescribe pain medication**? | Can you give an example where effects of pain meds influenced your decision?  Can you give an example where side effects influenced your decision? |  |
| Follow up Presents of other comorbidities (from page 9) | Comorbidities | How does comorbidities affect your decision to prescribe or not? | -Are there particular comorbidities that stops you more often than others?  Can you give an example where comorbidities influenced your decision? |  |
| Follow up page 9 (top) of questionnaire | Alternative strategies | Some research studies have tried to investigate the use of alternative pain management strategies (f.ex. change in physical activity) instead of using pain medication. **Have you tried alternative treatments?** | ~~-~~ What would you think an effective alternative pain management strategy might be? |  |
| Follow up Knowledge of the patient (from page 9 | Knowledge of the patient | In the questionnaire, a proposed factor that might influence the prescription of pain medication was the **relationship/knowledge with/of the patient.** This includes the patient’s **previous treatment history, the patient’s expectations** and the patient’s health literacy. **Does this also relate to you?** | Can you give an example where these factors have made you do one thing or the other? |  |
| Follow up Health literacy | Health literacy | Next, I would like to ask you about the term sundhedskompetencer, aka health literacy. It is a rather new term in Denmark, and is understood as a person’s ability to access, understand, appraise and implement health information in their daily life, in order to increase QoL, prevent diseases and promote health.  Is this something you consider to be a part of your meeting with a MSK pain patient? Directly or indirectly.  (They may not have time to really take it into consideration).  How so? | How do they estimate HL?  How do they react/respond if they find a patient has limited or poor HL?  Do they ensure that patients understand? If so, how?  Do they see a tendency regarding what type of patients generally seem to have low HL? |  |
| Rounding up the interview | (Additional response) | Is there something we have not covered, and/or you would like to add? |  |  |
| Finishing off |  | Thank you for your participation!  (Feel free to contact us if you have further questions) |  |  |
