## Appendix 2 for "How do general practitioners consider health literacy in pain medication treatment of patients suffering from chronic musculoskeletal pain? *a mixed methods study*"

**Appendix 2 – Literature Search**

**Databases and results**

| Database | Platform | Results | Date |
| --- | --- | --- | --- |
| PubMed | PubMed.gov | 7290 | 10.07.2025 |
| Embase | Embase.com | 8795 | 10.07.2025 |
| PsycINFO | APA | 10 | 10.07.2025 |
| All  ÷ duplicates with Covidence |  | 16095  1286 |  |

**Pubmed**

**Search 1: Health literacy + General practice + Chronic MSK pain + Pain medication**

| Search | Query | Results |
| --- | --- | --- |
| #1 | Search: ("general practitioner*" OR "family physician*" OR "primary care physician*" OR "GP*" OR "primary care") AND ("chronic musculoskeletal pain" OR "chronic MSK pain" OR "chronic pain" OR "musculoskeletal disorder*") AND ("health literacy" OR "health communication" OR "patient understanding" OR "health knowledge" OR "health information seeking") AND ("pain medication" OR "analgesics" OR "opioid*" OR "NSAID*" OR "prescription" OR "pharmacological treatment") AND ("adult*" OR "adults" OR "middle aged" OR "older adults") | 25 |

**Search 2: Health literacy + General practice + Chronic MSK pain**

| Search | Query | Results |
| --- | --- | --- |
| #2 | Search: ("general practitioner*" OR "family physician*" OR "primary care physician*" OR "GP*" OR "primary care") AND ("chronic musculoskeletal pain" OR "chronic MSK pain" OR "chronic pain" OR "musculoskeletal disorder*") AND ("health literacy" OR "health communication" OR "patient understanding" OR "health knowledge" OR "health information seeking") AND ("adult*" OR "adults" OR "middle aged" OR "older adults") | 83 |

**Search 3: Health literacy + General practice**

| Search | Query | Results |
| --- | --- | --- |
| #3 | Search: ("general practitioner*" OR "family physician*" OR "primary care physician*" OR "GP*" OR "primary care") AND ("health literacy" OR "health communication" OR "patient understanding" OR "health knowledge" OR "health information seeking") AND ("adult*" OR "adults" OR "middle aged" OR "older adults") | 7182 |

**Embase**

**Search 1: Health literacy + General practice + Chronic MSK pain + Pain medication**

| Search | Query | Results |
| --- | --- | --- |
|  | Search: ('general practitioner'/exp OR 'family physician'/exp OR 'primary medical care'/exp OR 'general practitioner*':ti,ab OR 'family physician*':ti,ab OR 'primary care':ti,ab) AND ('musculoskeletal pain'/exp OR 'chronic pain'/exp OR 'musculoskeletal disease'/exp OR 'chronic musculoskeletal pain':ti,ab OR 'chronic pain':ti,ab) AND ('health literacy'/exp OR 'health communication'/exp OR 'patient education'/exp OR 'health knowledge':ti,ab OR 'health literacy':ti,ab OR 'patient understanding':ti,ab) AND ('analgesic agent'/exp OR 'opioid'/exp OR 'nonsteroidal antiinflammatory agent'/exp OR 'pain medication':ti,ab OR 'NSAID*':ti,ab OR 'opioid*':ti,ab OR 'prescription':ti,ab) AND ('adult'/exp OR 'adult*':ti,ab OR 'middle aged':ti,ab) | 287 |

**Search 2: Health literacy + General practice + Chronic MSK pain**

| Search | Query | Results |
| --- | --- | --- |
| #1 | Search: ('general practitioner'/exp OR 'primary medical care'/exp OR 'general practitioner*':ti,ab OR 'primary care':ti,ab) AND ('musculoskeletal pain'/exp OR 'chronic pain'/exp OR 'chronic musculoskeletal pain':ti,ab) AND ('health literacy'/exp OR 'health communication'/exp OR 'patient education'/exp OR 'health literacy':ti,ab OR 'patient understanding':ti,ab) AND ('adult'/exp OR 'adult*':ti,ab) | 295 |

**Search 3: Health literacy + General practice**

| Search | Query | Results |
| --- | --- | --- |
| #1 | Search: ('general practitioner'/exp OR 'primary medical care'/exp OR 'general practitioner*':ti,ab OR 'primary care':ti,ab) AND ('health literacy'/exp OR 'health communication'/exp OR 'patient education'/exp OR 'health literacy':ti,ab OR 'patient understanding':ti,ab) AND ('adult'/exp OR 'adult*':ti,ab) | 8213 |

**PsycINFO**

**Search 1: Health literacy + General practice + Chronic MSK pain + Pain medication**

| Search | Query | Results |
| --- | --- | --- |
| #1 | Search: (DE "General Practitioners" OR DE "Primary Health Care" OR "general practitioner*":ti,ab OR "primary care":ti,ab) AND (DE "Chronic Pain" OR DE "Musculoskeletal System" OR "chronic musculoskeletal pain":ti,ab OR "musculoskeletal disorder*":ti,ab) AND (DE "Health Literacy" OR DE "Health Communication" OR DE "Patient Education" OR "health literacy":ti,ab OR "health knowledge":ti,ab OR "patient understanding":ti,ab) AND (DE "Drug Therapy" OR DE "Analgesics" OR "pain medication":ti,ab OR "NSAID*":ti,ab OR "opioid*":ti,ab) AND (DE "Adults" OR "adult*":ti,ab OR "middle aged":ti,ab) | 1 |

**Search 2: Health literacy + General practice + Chronic MSK pain**

| Search | Query | Results |
| --- | --- | --- |
| #1 | Search: (DE "General Practitioners" OR DE "Primary Health Care" OR "general practitioner*":ti,ab OR "primary care":ti,ab) AND (DE "Chronic Pain" OR DE "Musculoskeletal System" OR "chronic musculoskeletal pain":ti,ab) AND (DE "Health Literacy" OR DE "Health Communication" OR DE "Patient Education" OR "health literacy":ti,ab) AND (DE "Adults" OR "adult*":ti,ab) | 1 |

**Search 3: Health literacy + General practice**

| Search | Query | Results |
| --- | --- | --- |
| #1 | Search: (DE "General Practitioners" OR DE "Primary Health Care" OR "general practitioner*":ti,ab OR "primary care":ti,ab) AND (DE "Health Literacy" OR DE "Health Communication" OR DE "Patient Education" OR "health literacy":ti,ab) AND (DE "Adults" OR "adult*":ti,ab) | 8 |
